# Impact of Physician Experience on Stroke or Death Rates in Transfemoral Carotid Artery Stenting: Insights from the Vascular Quality Initiative

**DOI:** 10.1101/2023.11.16.23298660

**Authors:** Gabriel Jabbour, Sai Divya Yadavalli, Sabrina Strauss, Andrew P. Sanders, Vinamr Rastogi, Jens Eldrup-Jorgensen, Richard J. Powell, Roger B. Davis, Marc L. Schermerhorn

## Abstract

**Objective:** With the recent expansion of the Centers for Medicare and Medicaid Services (CMS) coverage, transfemoral carotid artery stenting (tfCAS) is expected to play a larger role in the management of carotid disease. Existing research on the tfCAS learning curve, primarily conducted over a decade ago, may not adequately describe the current effect of physician experience on outcomes. This study evaluates the tfCAS learning curve using VQI data.

**Methods:** We analyzed tfCAS patient data from 2005-2023. Each physician’s procedures were chronologically grouped into 12 categories, from procedure counts 1-25 to 351+. Primary outcome was in-hospital stroke/death rate; secondary outcomes were in-hospital stroke/death/MI, 30-day mortality, and in-hospital stroke/TIA. The relationship between outcomes and procedure counts was analyzed using Cochran Armitage test and a generalized linear model with restricted cubic splines, validated using generalized estimating equations.

**Results:** We analyzed 43,147 procedures by 2,476 physicians. In symptomatic patients, there was a decrease in rates of in-hospital stroke/death (procedure counts 1-25 to 351+: 5.2% to 1.7%), in-hospital stroke/death/MI (5.8% to 1.7%), 30-day mortality (4.6% to 2.8%), in-hospital stroke/TIA (5.0% to 1.1%) (all p-values<0.05). The in-hospital stroke/death rate remained above 4% until 235 procedures. Similarly, in asymptomatic patients, there was a decrease in rates of in-hospital stroke/death (2.1% to 1.6%), in-hospital stroke/death/MI (2.6% to 1.6%), 30-day mortality (1.7% to 0.4%), and in-hospital stroke/TIA (2.8% to 1.6%) with increasing physician experience (all p-values<0.05). The in-hospital stroke/death rate remained above 2% until 13 procedures.

**Conclusions:** In-hospital stroke/death and 30-day mortality rates post-tfCAS decreased with increasing physician experience, showing a lengthy learning curve consistent with previous reports. Given that physicians’ early cases may not be included in the VQI, the learning curve was likely underestimated. With the recent CMS coverage expansion for tfCAS, a significant number of physicians would enter the early stage of the learning curve, potentially leading to increased post-operative complications.

**ARTICLE HIGHLIGHTS:** *Type of Research:* Retrospective analysis of prospectively collected Vascular Quality Initiative registry data.

*Key Findings:* In patients undergoing tfCAS in VQI, in-hospital stroke/death, in-hospital stroke/death/MI, 30-day mortality and in-hospital stroke/TIA decreased with increasing physician experience in both symptomatic and asymptomatic patients. In symptomatic patients, in-hospital stroke/death rate did not drop below 4% until after 235 procedures, and it remained above 2% until 13 procedures in asymptomatic patients.

*Take home Message:* This study showed a decrease in post-operative in-hospital stroke/death with a substantially high risk in an operator’s first 25 procedures in VQI. The recent expansion of the Center for Medicare and Medicare Services coverage of tfCAS warrants caution since a rise in early-phase physicians could lead to increased post-operative complication rates in transfemoral carotid artery stent patients.

*Table of Contents Summary:* In this retrospective analysis of the tfCAS learning curve, in-hospital stroke/death, in-hospital stroke/death/MI, 30-day mortality, and in-hospital stroke/TIA rates decreased significantly with increased physician experience. With the recent CMS coverage expansion for tfCAS, more physicians would enter the early stage of the learning curve, potentially leading to increased post-operative complications.

## Introduction

Despite its minimally invasive nature and two decades of technological progress, transfemoral carotid artery stenting (tfCAS) has not developed a clearly defined role in the treatment of carotid stenosis.^1,2,3^ Previous studies have shown higher periprocedural stroke rates after tfCAS compared with carotid endarterectomy (CEA), which has impacted the reimbursement policies in the United States.^1,2,3,4^ Similarly, transcarotid artery revascularization (TCAR) has been shown to have lower perioperative stroke/death rate compared with tfCAS^5,6,7^ and half the risk of stroke/death/TIA despite patients having more comorbidities.^7^ Even in patients at high risk for CEA, carotid stenting has not been clearly demonstrated to be advantageous.^8,9^

Recently, the Multispecialty Carotid Alliance sent a formal request to the Centers for Medicare and Medicaid Services (CMS) to expand their coverage for tfCAS beyond patients at high risk for CEA^10^. In response, the CMS announced its final decision to expand coverage for tfCAS, which would allow a larger number of patients and physicians to utilize this treatment option.^11,12^ However, broadening access to tfCAS might have adverse consequences. TfCAS is an advanced endovascular technique and has been shown to have a lengthy learning curve, which could lead to variations in outcomes as practitioners strive to become proficient in the procedure. Multiple analyses of the tfCAS learning curve have demonstrated that initial procedures have higher complication rates putting patients at risk for stroke.^13,14,15^ These findings underscore the value of physician experience with the procedure and emphasize the significant learning curve associated with this advanced endovascular procedure.

Despite the wealth of studies on carotid artery stenting, there has been no recent analysis of the learning curve of tfCAS. Research on the tfCAS learning curve was primarily conducted over a decade ago and may not adequately reflect current physician experience.^13,14,15,16,17^ Furthermore, prior studies did not account for the likelihood that early successes might motivate physicians to continue performing the procedure more frequently, ultimately leading to higher procedure counts. Some of those studies had previously shown a linear relationship between increasing physician experience and decreasing peri-operative complications.^14,17^ Following the CMS coverage expansion, the number of patients undergoing tfCAS is expected to increase and more novice physicians may begin performing this procedure leading to an increased rate of post-operative complications. Since about 25-30% of stroke/death post-CAS occur after discharge, ^18,19,20^ appropriate thresholds for in-hospital event rates (stroke/death) have been suggested to be 4% for symptomatic and 2% for asymptomatic patients instead of 6% for symptomatic and 3% for asymptomatic 30-day event rates.^21,22^ As the VQI has the most complete data for in-hospital outcomes, we have chosen this time period as the basis for the study using 4% for symptomatic and 2% for asymptomatic patients as cutoffs for the acceptable in-hospital event rates.

In this manuscript, we aim to evaluate the tfCAS learning curve based on the Vascular Quality Initiative (VQI) database to better understand the potential risks and challenges associated with tfCAS in the hands of less experienced operators.

## Methods

### Data Source

We used data from the Carotid Artery Stenting (CAS) registry of the VQI, which is a clinical registry supplied by prospectively collected data from the United States, Canada and Singapore. The registry collects demographic, clinical, procedural and outcomes data to improve the quality, safety, effectiveness, and cost of vascular procedures. The institutional review board at Beth Israel Deaconess Medical Center has approved the use of deidentified data from the VQI for research purposes with no need for informed consent.

### Patient cohort and variable definitions

All patients who underwent tfCAS from March 2005 through June 2023 were included in the study. Procedures performed by each physician were numbered in chronological order as procedure count irrespective of symptom status. For each procedure count, physician count was calculated as the total number of physicians who have performed at least that many procedures without discriminating between symptomatic or asymptomatic patients. Procedure categories were created for procedure counts 1-25, 26-50, 51-75, 76-100, 101-125, 126-150, 151-175, 176-200, 201-250, 251-300, 301-350, and 351+.

Patients with an ipsilateral neurologic symptom within 180 days prior to their procedure were considered symptomatic. Chronic kidney disease (CKD) was defined as having an estimated glomerular filtration rate (eGFR) < 60 ml/min, which was further categorized into eGFR 45-59.9, eGFR 30-44.9, and eGFR < 30 or being on hemodialysis.^23,24,25^

The primary outcome of our study was in-hospital stroke or death (stroke/death) rate. Secondary outcomes were in-hospital stroke/death/MI, in-hospital stroke/death/TIA, in-hospital death, in-hospital stroke, 30-day mortality, post-operative neurologic events (stroke/TIA), and access site complications.

### Statistical Analysis

Categorical variables were presented as percentages. Continuous variables were presented as mean (standard deviation) given they were normally distributed. Missing values were not dropped and were shown in the tables only if they exceeded 5% per variable. For bivariable analyses, the Cochran Armitage test was used to study trends in binary variables in different procedure categories. P-values <.05 were considered statistically significant.

We employed a generalized linear model (GLM) to analyze the unadjusted relationship between the outcome variable and the procedure count, incorporating the use of restricted cubic splines (RCS) to allow for potential non-linearities. Knots are predefined values where the spline’s slope can change, allowing the model to capture complex relationships between predictor and response variables. The number of knots for the RCS was determined for each outcome variable using the Akaike information criterion (AIC) to achieve a balance between flexibility and overfitting. To account for the possibility that physicians with better early outcomes might end up performing more procedures, we validated our results with a sensitivity analysis using a generalized estimating equation (GEE) model clustered by physician ID. All analyses were performed using R version 4.3.1 (R Core Team, 2023).

## Results

A total of 43,147 procedures performed by 2476 unique physicians were included in the analysis, 39% being performed on symptomatic patients and 61% on asymptomatic patients.

### Symptomatic patients

#### Baseline characteristics

Baseline characteristics of symptomatic patients undergoing tfCAS stratified by physician procedure count category can be found in Table 1A. As physician experience increased, patients were more likely to be greater than 75 years old (p=.006) and a less likely to identify as non-white (p=.002), have hypertension (HTN) (p=.032), coronary artery disease (CAD) (p<.001), moderate or severe anemia (p=.036), undergo an urgent/emergent procedure (p=.002), present with ipsilateral carotid stenosis ≥ 80% (p=.002), and be anatomically high risk for CEA (p<.001) (Table 1A).

**Table 1A.**
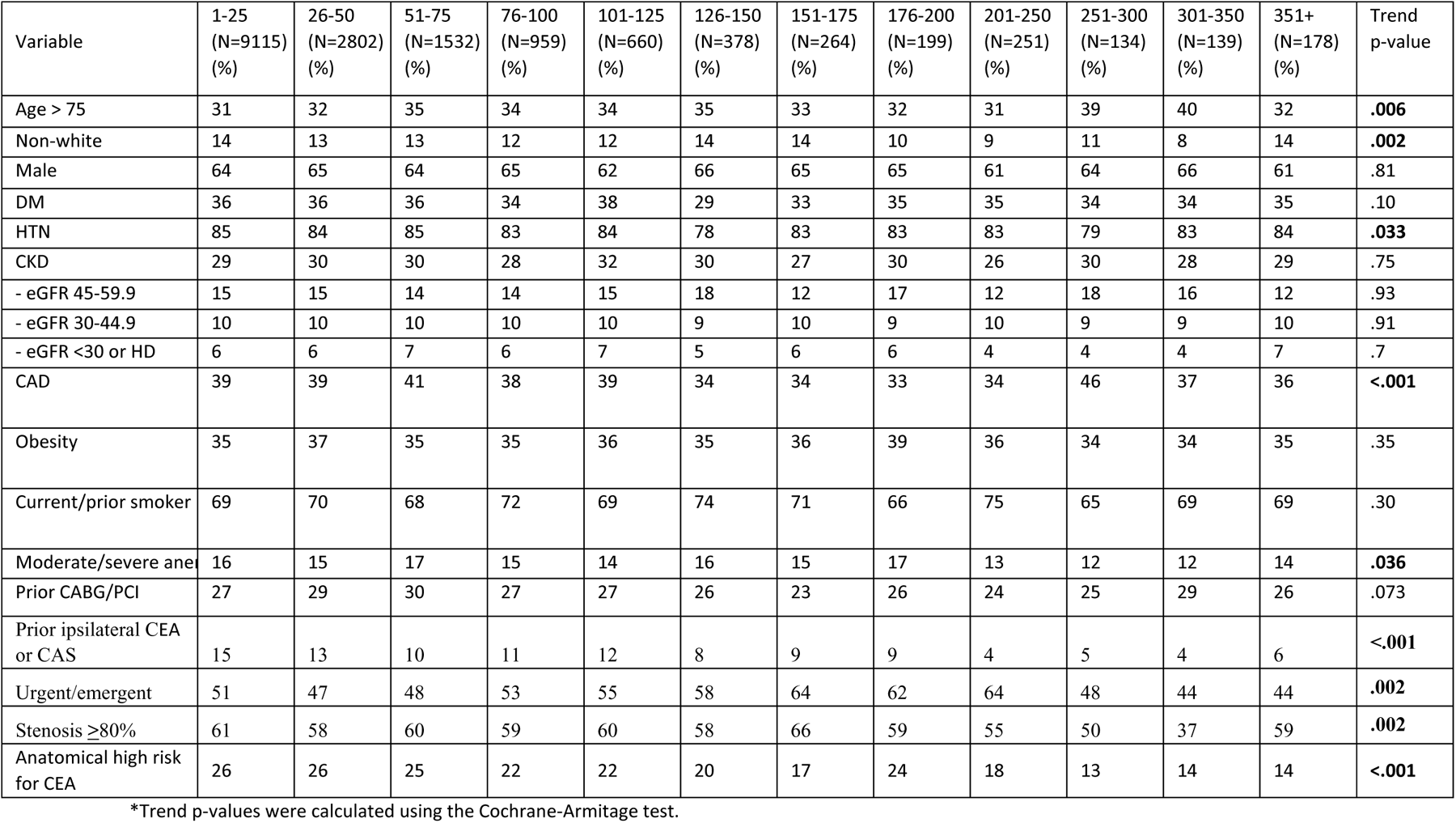
Baseline characteristics of symptomatic patients who underwent tfCAS stratified by procedure count category.

**Table 1B.** Baseline characteristics of asymptomatic patients who underwent tfCAS stratified by procedure count category.

| Variable | 1-25<br>(N=15788)<br>(%) | 26-50<br>(N=4146)<br>(%) | 51-75<br>(N=2144)<br>(%) | 76-100<br>(N=1226)<br>(%) | 101-125<br>(N=792)<br>(%) | 126-150<br>(N=498)<br>(%) | 151-175<br>(N=356)<br>(%) | 176-200<br>(N=270)<br>(%) | 201-250<br>(N=410)<br>(%) | 251-300<br>(N=324)<br>(%) | 301-350<br>(N=217)<br>(%) | 351+<br>(N=250)<br>(%) | Trend<br>p-value |
| --- | --- | --- | --- | --- | --- | --- | --- | --- | --- | --- | --- | --- | --- |
| Age > 75 | 32 | 33 | 35 | 34 | 35 | 30 | 30 | 33 | 36 | 37 | 36 | 34 | **.039** |
| Non-white | 11 | 10 | 9 | 9 | 9 | 9 | 9 | 7 | 8 | 8 | 6 | 8 | **<.001** |
| Male | 63 | 64 | 64 | 63 | 62 | 61 | 62 | 64 | 62 | 65 | 67 | 62 | .859 |
| DM | 38 | 38 | 39 | 36 | 35 | 40 | 37 | 41 | 40 | 41 | 36 | 42 | .48 |
| HTN | 88 | 89 | 88 | 87 | 86 | 87 | 82 | 86 | 88 | 88 | 89 | 90 | .70 |
| CKD | 35 | 36 | 37 | 35 | 37 | 37 | 30 | 40 | 34 | 34 | 36 | 41 | .47 |
| - eGFR 45-59.9 | 18 | 18 | 18 | 18 | 19 | 18 | 17 | 17 | 19 | 17 | 18 | 17 | .42 |
| - eGFR 30-44.9 | 12 | 13 | 13 | 11 | 13 | 12 | 7 | 19 | 9 | 13 | 10 | 15 | .8 |
| - eGFR <30 or HD | 7 | 7 | 7 | 8 | 6 | 7 | 8 | 6 | 7 | 4 | 11 | 12 | .11 |
| CAD | 51 | 51 | 51 | 52 | 50 | 48 | 48 | 52 | 47 | 55 | 50 | 57 | **<.001** |
| Obesity | 34 | 34 | 35 | 37 | 35 | 36 | 38 | 40 | 36 | 36 | 32 | 36 | **.005** |
| Current/prior smoker | 73 | 74 | 75 | 75 | 72 | 76 | 71 | 75 | 80 | 71 | 80 | 76 | **.016** |
| Moderate/severe anemia | 16 | 15 | 17 | 16 | 15 | 17 | 14 | 18 | 13 | 15 | 14 | 14 | .31 |
| Prior CABG/PCI | 37 | 38 | 38 | 38 | 36 | 34 | 36 | 37 | 34 | 43 | 36 | 44 | .28 |
| Prior ipsilateral CEA or CAS | 26 | 23 | 20 | 21 | 17 | 17 | 19 | 18 | 16 | 9 | 11 | 11 | **<.001** |
| Urgent/emergent | 16 | 16 | 16 | 16 | 14 | 15 | 11 | 15 | 12 | 9 | 8 | 6 | **<.001** |
| Stenosis ≥80% | 56 | 56 | 55 | 54 | 57 | 55 | 54 | 52 | 42 | 40 | 47 | 57 | **<.001** |
| Anatomical High Risk for CEA | 29 | 29 | 27 | 27 | 23 | 28 | 28 | 27 | 23 | 15 | 13 | 22 | **<.001** |

\*Trend p-values were calculated using the Cochrane-Armitage test.

#### Procedural characteristics

In symptomatic patients, increased physician experience was associated with a decrease in total procedure time (p<.001), decrease in total fluoroscopy time (p<.001), protamine use (p<.001); and an increase in the use of distal embolic protection device (p<.001) (Table 2A). However, the dataset contains a high rate of missing values, which were included in the table.

**Table 2A.** Procedural characteristics in symptomatic patients who underwent tfCAS stratified by procedure count category.

| Variable | 1-25<br>(N=9115) | 26-50<br>(N=2802) | 51-75<br>(N=1532) | 76-100<br>(N=959) | 101-125<br>(N=660) | 126-150<br>(N=378) | 151-175<br>(N=264) | 176-200<br>(N=199) | 201-250<br>(N=251) | 251-300<br>(N=134) | 301-350<br>(N=139) | 351+<br>(N=178) | Trend<br>p-value* |
| --- | --- | --- | --- | --- | --- | --- | --- | --- | --- | --- | --- | --- | --- |
| Total procedure time:<br>mean (SD) | 84 (48) | 79 (54) | 77 (46) | 68 (45) | 69 (50) | 73 (47) | 86 (57) | 64 (36) | 58 (25) | 55 (23) | 56 (24) | 65 (27) | **<.001** |
| - Missing | 34% | 28% | 28% | 21% | 17% | 25% | 20% | 18% | 13% | 10% | 6% | 3% |  |
| Fluoro time: mean (SD) | 26 (30) | 24 (20) | 26 (24) | 24 (20) | 24 (25) | 24 (22) | 27 (20) | 21 (18) | 21 (24) | 21 (20) | 21 (24) | 12 (9) | **<.001** |
| - Missing | 36% | 29% | 29% | 21% | 17% | 25% | 20% | 21% | 13% | 10% | 5% | 0% |  |
| Contrast volume:<br>mean (SD) | 102 (58) | 103 (60) | 108 (61) | 105 (62) | 109 (63) | 110 (63) | 105 (63) | 106 (61) | 101 (67) | 122 (83) | 120 (62) | 68 (45) | .24 |
| - Missing | 5% | 3% | 2% | 3% | 4% | 2% | 3% | 1% | 0% | 0% | 0% | 1% |  |
| Protamine use (%) | 13% | 13% | 12% | 12% | 15% | 15% | 17% | 7% | 9% | 2% | 2% | 2% | **<.001** |
| - Missing | 12% | 15% | 13% | 11% | 9% | 10% | 16% | 17% | 12% | 23% | 28% | 22% |  |
| Distal Embolic Protection<br>Device Use (%) | 84% | 86% | 87% | 88% | 84% | 83% | 72% | 83% | 87% | 85% | 86% | 93% | **<.001** |
| - Missing | 14% | 11% | 9% | 8% | 12% | 11% | 22% | 13% | 10% | 13% | 14% | 7% |  |

\*Trend p-values were calculated using the Cochran-Armitage test for categorical variables and the Jonckheere Terpstra test for continuous variables

**Table 2B.**
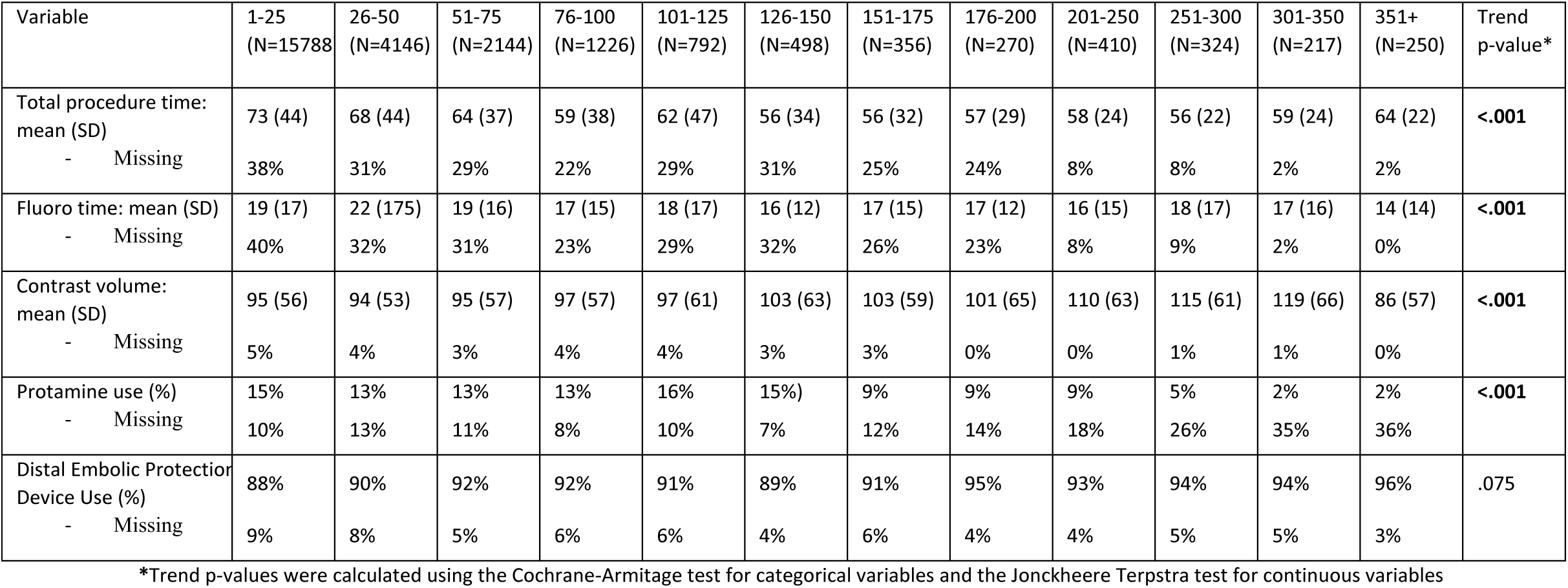
Procedural characteristics in asymptomatic patients who underwent tfCAS stratified by procedure count category.

#### Post-procedural outcomes

In symptomatic patients, a decrease was seen with increased physician experience for in-hospital stroke/death rate (procedure count 1-25 to 351+: 5.2% to 1.7%, p=.036), in-hospital stroke/death/MI rate (5.8% to 1.7%, p=.039), in-hospital stroke/death/TIA rate (6.3% to 1.7%, p=.009), in-hospital stroke (2.6% to 1.1%, p=.072), 30-day mortality (4.6% to 2.8%, p=.042), post-operative neurologic event rate (5.0% to 1.1%, p<.001), and access site complication rate (4.1% to 1.1%, p=.002). Moreover, there was a trend toward a decrease in in-hospital death (3.3% to 0.6%, p=.074) and in-hospital stroke (2.6% to 1.1%, p=.072) (Table 3A). However, there were substantial variations in all outcomes variables and the incidence of events frequently went back above thresholds despite significant physician experience.

**Table 3A.** Post-operative outcomes in symptomatic patients who underwent tfCAS stratified by procedure count category.

| Variable | 1-25<br>(N=9115)<br>(%) | 26-50<br>(N=2802)<br>(%) | 51-75<br>(N=1532)<br>(%) | 76-100<br>(N=959)<br>(%) | 101-125<br>(N=660)<br>(%) | 126-150<br>(N=378)<br>(%) | 151-175<br>(N=264)<br>(%) | 176-200<br>(N=199)<br>(%) | 201-250<br>(N=251)<br>(%) | 251-300<br>(N=134)<br>(%) | 301-350<br>(N=139)<br>(%) | 351+<br>(N=178)<br>(%) | Trend<br>p-value* |
| --- | --- | --- | --- | --- | --- | --- | --- | --- | --- | --- | --- | --- | --- |
| In-hospital stroke/death | 5.2 | 4.8 | 5 | 5.2 | 3.9 | 5.6 | 4.9 | 4 | 3.2 | 6.7 | 3.6 | 1.7 | **.036** |
| In-hospital stroke/death/MI | 5.8 | 5.3 | 5.6 | 5.5 | 4.4 | 6.9 | 5.7 | 4.5 | 4 | 6.7 | 4.3 | 1.7 | **.039** |
| In-hospital stroke/death/TIA | 6.3 | 5.9 | 5.7 | 5.8 | 5 | 5.8 | 5.3 | 6.5 | 4 | 8.2 | 3.6 | 1.7 | **.009** |
| In-hospital death | 3.3 | 2.9 | 3.1 | 3 | 2.7 | 3.4 | 3.8 | 3 | 1.6 | 4.5 | 1.4 | 0.6 | .074 |
| In-hospital stroke | 2.6 | 2.5 | 2.4 | 2.8 | 1.5 | 2.9 | 1.9 | 1.5 | 1.6 | 3 | 2.2 | 1.1 | .072 |
| 30-day mortality | 4.6 | 4.1 | 4.4 | 4.5 | 3.9 | 4 | 4.5 | 3 | 3.6 | 4.5 | 1.4 | 2.8 | **.042** |
| In-hospital stroke/TIA | 5 | 4.6 | 3.8 | 4.3 | 3.3 | 3.7 | 2.7 | 4 | 2.4 | 5.2 | 2.2 | 1.1 | **<.001** |
| Access site complication | 4.1 | 3.5 | 3.9 | 4 | 4.1 | 4.2 | 2.7 | 1.5 | 3.6 | 2.2 | 0 | 1.1 | **.002** |

\*Trend p-values were calculated using the Cochrane-Armitage test.

**Table 3B.** Post-operative outcomes in asymptomatic patients who underwent tfCAS stratified by procedure count category.

| Variable | 1-25<br>(N=15788)<br>(%) | 26-50<br>(N=4146)<br>(%) | 51-75<br>(N=2144)<br>(%) | 76-100<br>(N=1226)<br>(%) | 101-125<br>(N=792)<br>(%) | 126-150<br>(N=498)<br>(%) | 151-175<br>(N=356)<br>(%) | 176-200<br>(N=270)<br>(%) | 201-250<br>(N=410)<br>(%) | 251-300<br>(N=324)<br>(%) | 301-350<br>(N=217)<br>(%) | 351+<br>(N=250)<br>(%) | Trend<br>p-value* |
| --- | --- | --- | --- | --- | --- | --- | --- | --- | --- | --- | --- | --- | --- |
| In-hospital stroke/death | 2.1 | 1.5 | 2.1 | 1.9 | 2 | 1 | 1.1 | 3 | 0.7 | 0.9 | 2.3 | 1.6 | **.037** |
| In-hospital stroke/death/MI | 2.6 | 1.8 | 2.4 | 2.1 | 2.3 | 1.8 | 1.7 | 3.3 | 1.5 | 1.2 | 2.3 | 1.6 | **.017** |
| In-hospital stroke/death/TIA | 2.9 | 2.4 | 2.6 | 2.5 | 2.5 | 1.4 | 1.4 | 3 | 1.2 | 1.2 | 2.3 | 1.6 | **.001** |
| In-hospital death | 1 | 0.7 | 1.1 | 0.7 | 1 | 0.2 | 0.3 | 2.2 | 0.5 | 0 | 1.4 | 0 | .062 |
| In-hospital stroke | 1.3 | 1 | 1.1 | 1.3 | 1 | 0.8 | 0.8 | 1.1 | 0.5 | 0.9 | 0.9 | 1.6 | .11 |
| 30-day mortality | 1.7 | 1.3 | 1.4 | 1.1 | 1.6 | 1 | 1.1 | 2.6 | 1.2 | 0 | 1.8 | 0.4 | **.019** |
| In-hospital stroke/TIA | 2.8 | 2.4 | 2 | 2.3 | 1.6 | 1.6 | 2 | 1.1 | 1.2 | 1.2 | 1.4 | 1.6 | **<.001** |
| Access site complication | 3.4 | 2.7 | 3.2 | 3.1 | 2.7 | 3.8 | 3.7 | 2.6 | 2 | 4.6 | 4.1 | 2.4 | .51 |

\*Trend p-values were calculated using the Cochrane-Armitage test.

Using the GLM to model outcome rates vs procedure count on a continuous axis, in-hospital stroke/death rate decreased with increasing procedure count. In the first 25 cases, in-hospital stroke/death rate was above 4.7%, then plateaued at an unacceptably high rate of 5%, and did not decrease to under 4% until after 235 procedures, after which it continued to improve (Figure 1A). In-hospital stroke/death/MI rate decreased from 6.1% to 4% after 310 procedures, to a low of 2.7% after 521 procedures (figure 1B). In-hospital mortality decreased from 3.3% to 2% after 285 procedures, to its lowest point of 1% after 521 procedures (figure 1C).

**Figure 1.**
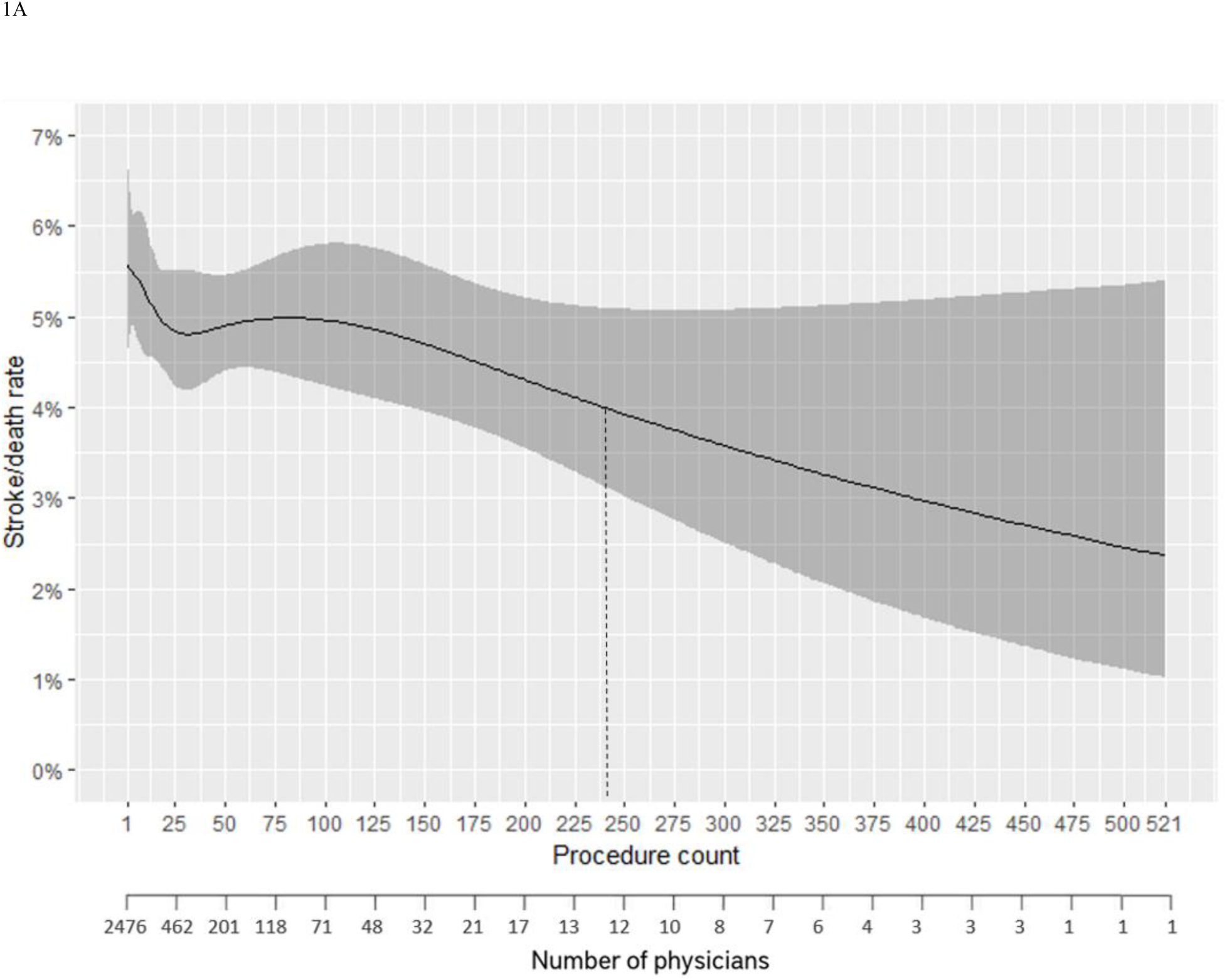
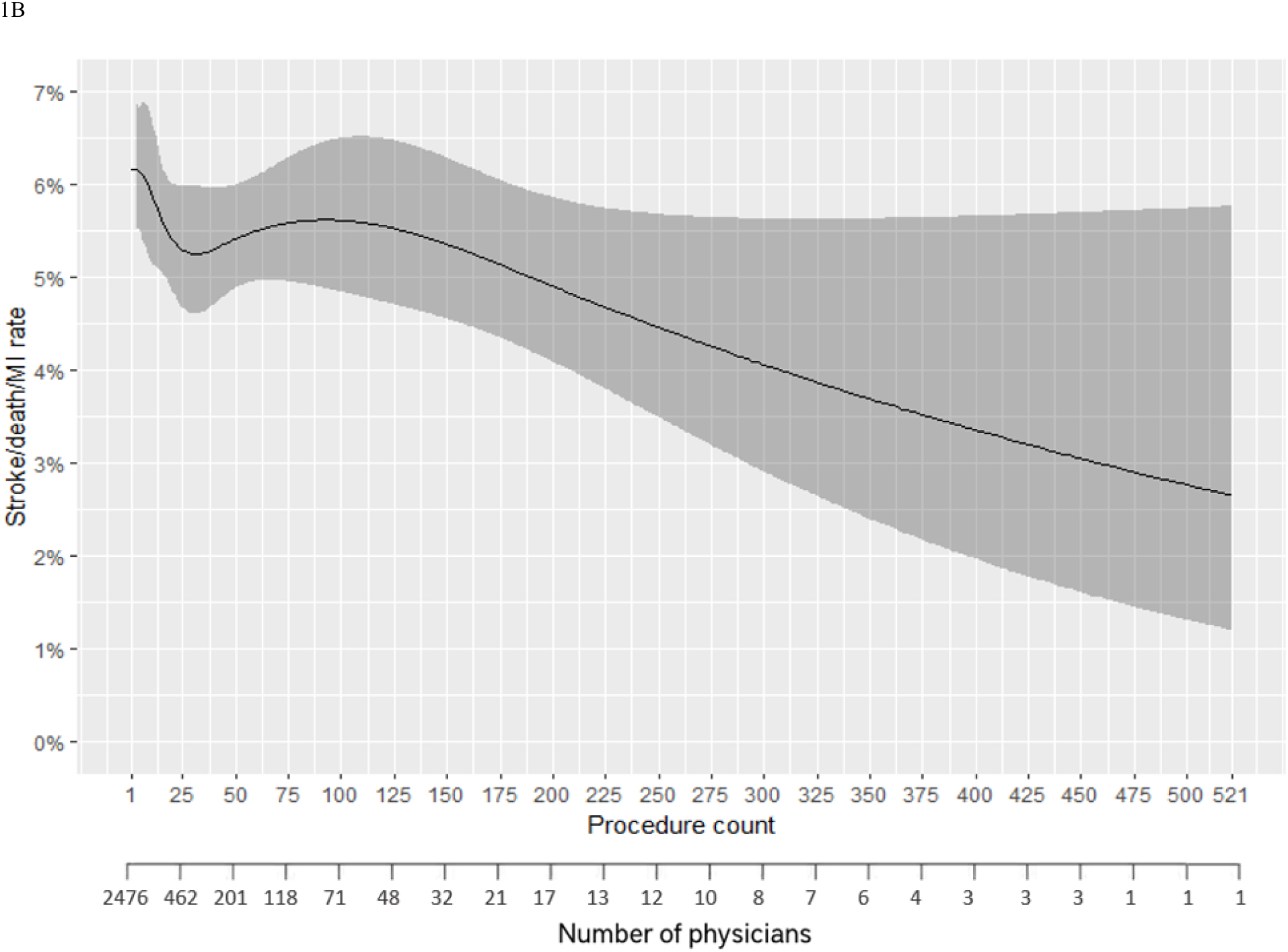
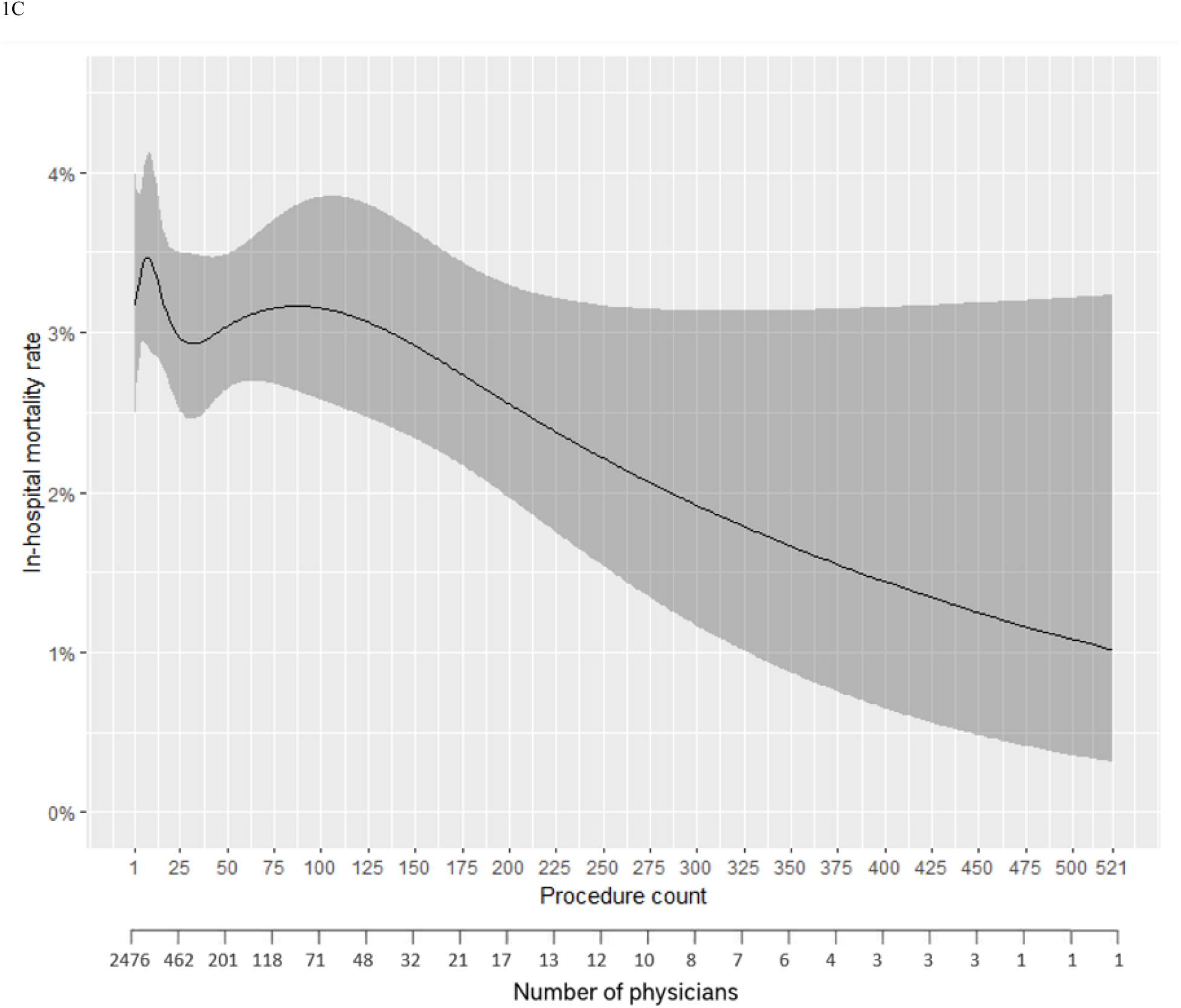
Generalized linear model of stroke/death (figure 1A), stroke/death/MI (figure 1B) and in-hospital mortality (figure 1C) of symptomatic patients who underwent tfCAS vs procedure count

Using the GEE model to adjust for the varying early outcomes between different physicians, an increase in physician experience was associated with a decrease in in-hospital stroke/death rate (p=.026), in-hospital stroke/death/MI (p=.03), and in-hospital mortality (p<.001).

### Asymptomatic patients

#### Baseline characteristics

In asymptomatic patients undergoing tfCAS, increased physician experience was associated with patients being greater than 75 years old (p=.039), obese (BMI ≥ 30) (p=.005), and current/prior smokers (p=.016). With increased physician experience, patients were less likely to identify as non-white (p<.001), have CAD (p<.001), have had prior ipsilateral CEA or CAS (p<.001), undergo an urgent/emergent procedure (p<.001), have ipsilateral carotid stenosis ≥ 80% (p<.001), and be anatomically high risk for CEA (p<.001) (Table 1B).

#### Procedural characteristics

In asymptomatic patients, increased physician experience was associated with a decrease in total procedure time (p<.001), decrease in total fluoroscopy time (p<.001), protamine use (p<.001); and an increase in the contrast volume used (p<.001) (Table 2B). However, the dataset contains a high rate of missing values, which were included in the table.

#### Post-procedural outcomes

In asymptomatic patients, a decrease was noted with increased physician experience for in-hospital stroke/death rate (2.1% to 1.6%, p=.037), in-hospital stroke/death/MI rate (2.6% to 1.6%, p=.017), in-hospital stroke/death/TIA rate (2.9% to 1.6%, p=.001), 30-day mortality (1.7% to 0.4%, p=.019), and post-operative neurologic event (2.8% to 1.6%, p<.001). In contrast, there was a trend toward a decrease in in-hospital death (1.0% to 0%, p=.062), but no significant change in in-hospital stroke (1.3% to 1.6%, p=.11) or access site complications (3.4% to 2.4%, p=.51) (Table 3B). However, there were substantial variations in all outcomes variables and the incidence of events frequently went back above thresholds despite substantial physician experience.

Using the GLM, in-hospital stroke/death rate decreased from 2.3% to 2% after 13 procedures, to 1% after 490 procedures, and kept decreasing (Figure 2A). In-hospital stroke/death/MI rate decreased from 2.6% to 2% after 210 procedures, to its lowest point of 1.1% at 526 procedures (Figure 2B). In-hospital mortality decreased from 1.1% to 1% after 10 procedures, to its lowest point of 0.4% after 526 procedures (Figure 2C).

**Figure 2.**
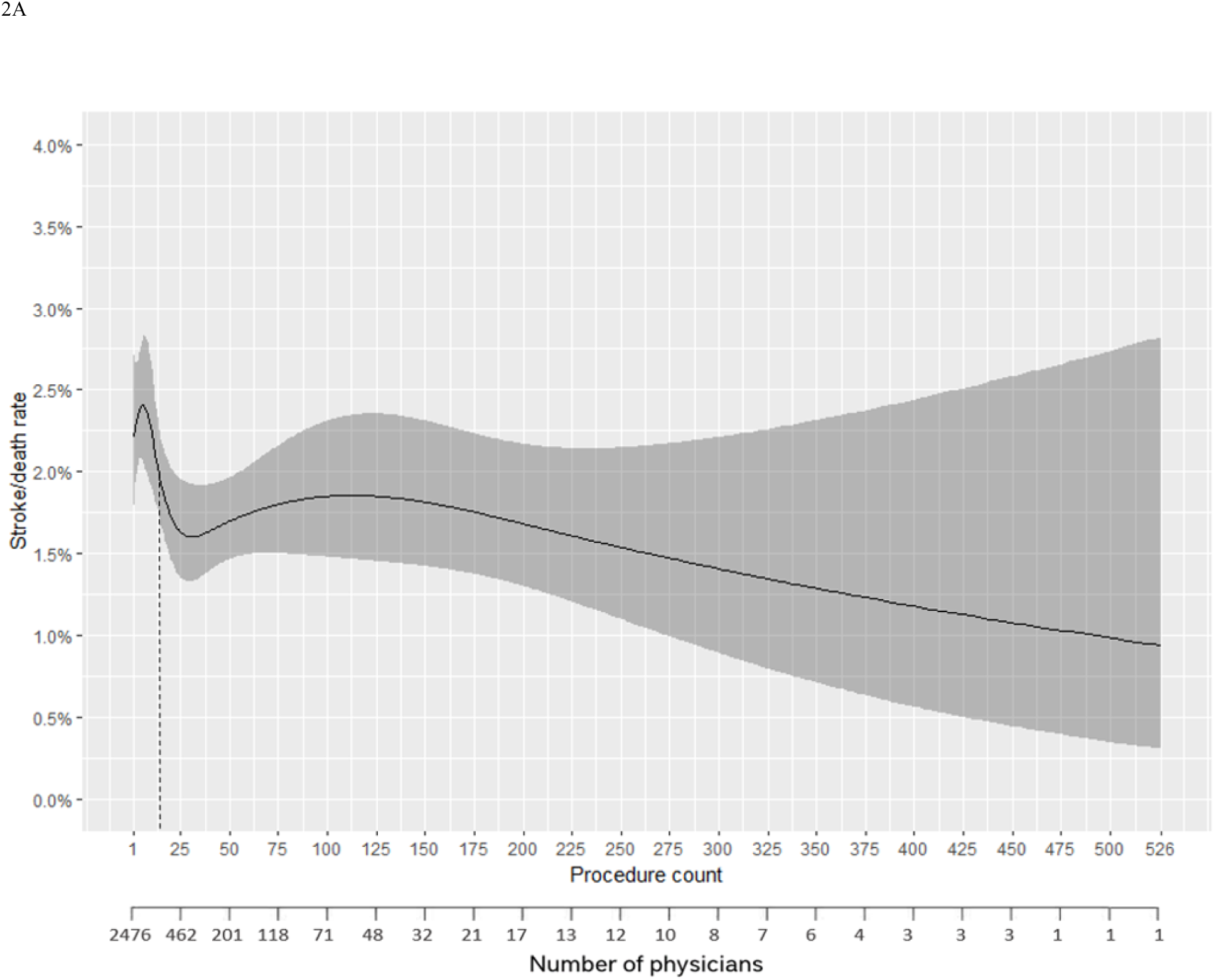
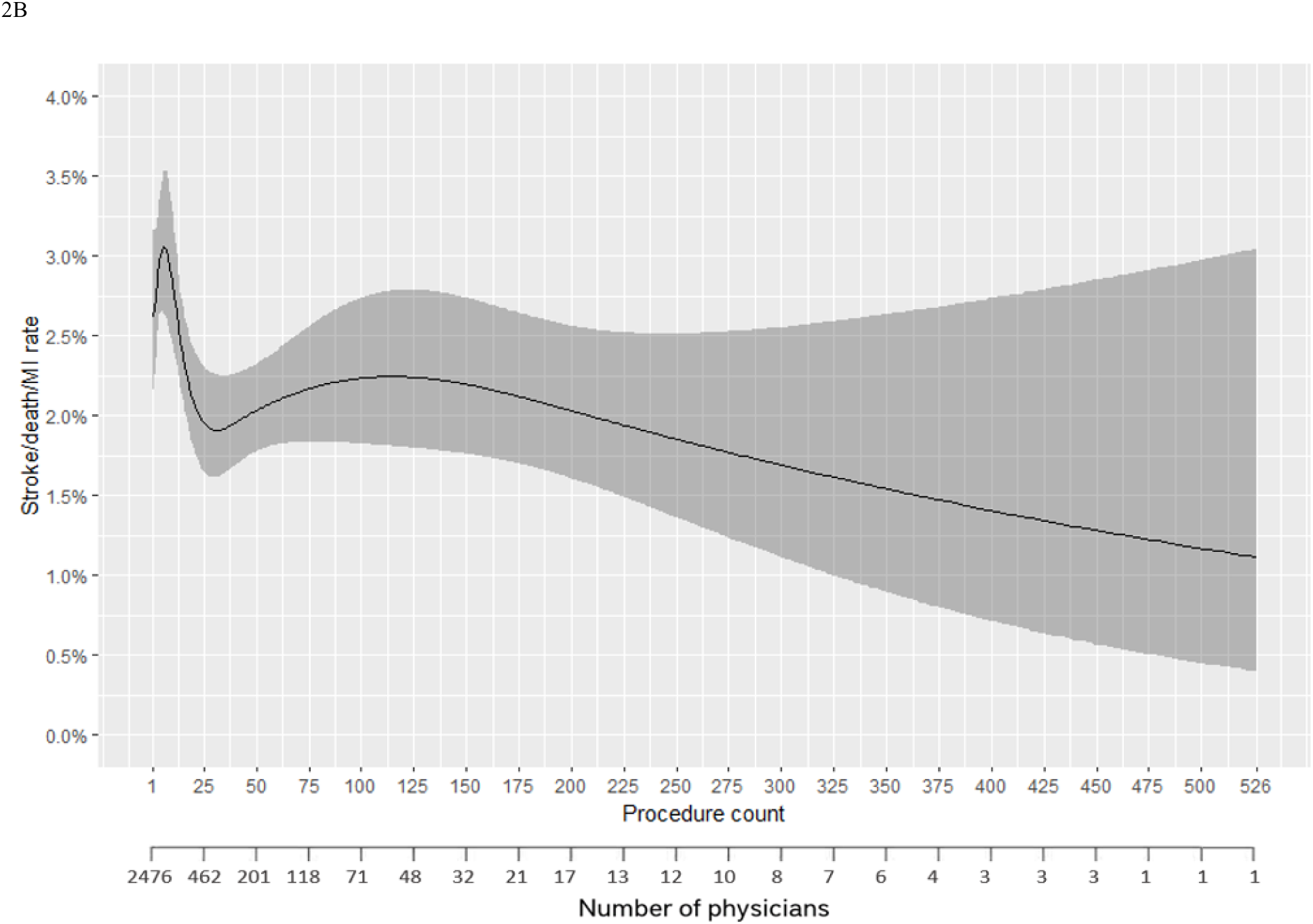
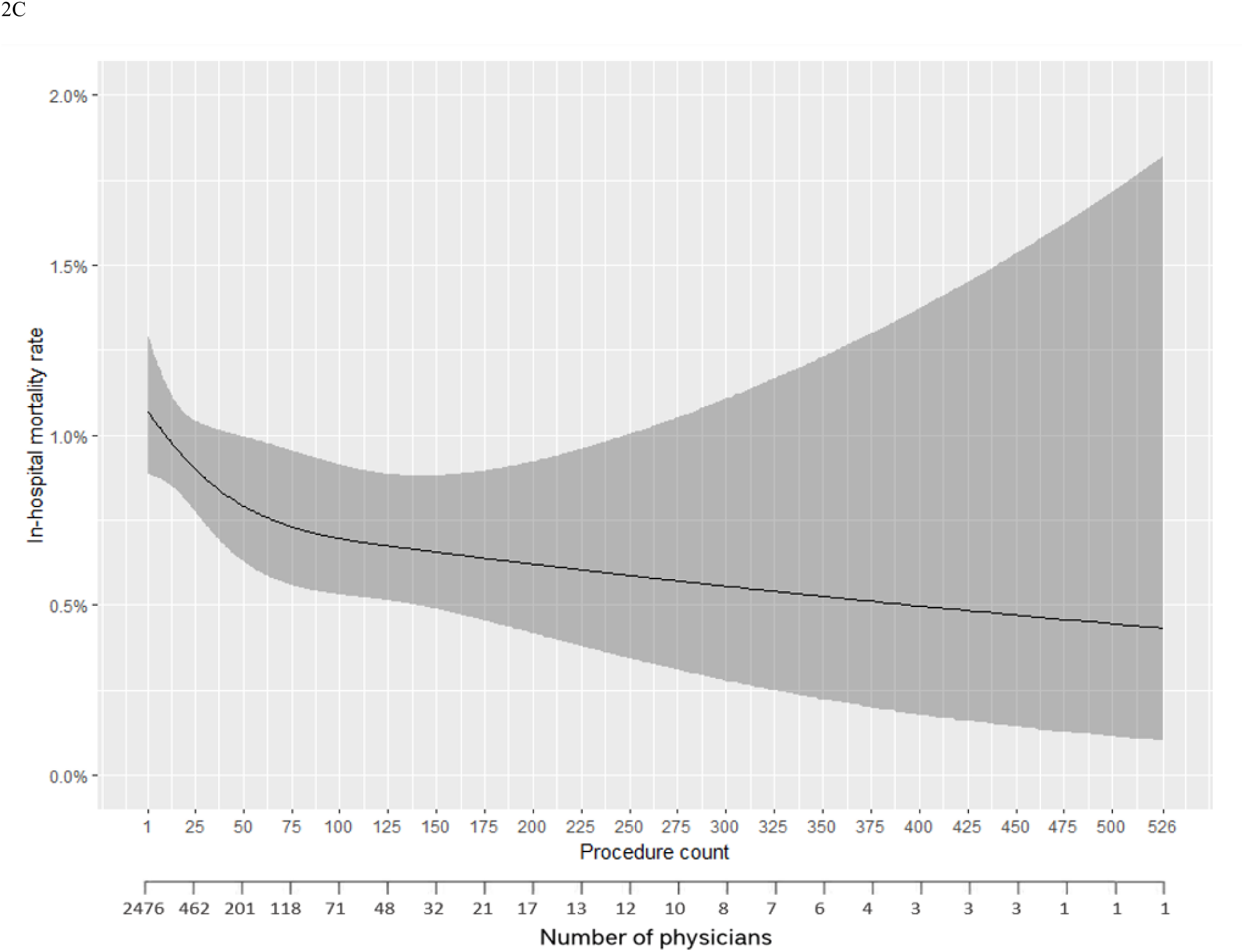
Generalized linear model of stroke/death (figure 2A), stroke/death/MI (figure 2B) and in-hospital mortality (figure 2C) of asymptomatic patients who underwent tfCAS vs procedure count

Using the GEE model to validate our previous results, an increase in physician experience was associated with a decrease in in-hospital stroke/death rate (p=.036), in-hospital stroke/death/MI (p=.013), and in-hospital mortality (p<.001).

### Sensitivity analysis

In a separate analysis, we explored the post-operative outcomes of physicians’ first 25 procedures as a factor of procedure count. In both symptomatic and asymptomatic patients, physicians who performed more than 25 procedures had lower rates of in-hospital stroke/death, in-hospital stroke/death/MI, in-hospital stroke/death/TIA, in-hospital death, and in-hospital stroke in their first 25 procedures compared to physicians who performed fewer than 25 procedures (p<.05) (Supplementary Tables S1A and S1B). Furthermore, we analyzed the post-operative outcomes for physicians who performed at least 25 procedures, stratified by procedure count category. For both symptomatic and asymptomatic patients, in-hospital stroke/death, in-hospital stroke/death/MI, in-hospital stroke/death/TIA, in-hospital death and in-hospital stroke rates did not change significantly with increasing physician experience (Supplementary Tables S2A and S2B). Nevertheless, the incidence of post-operative complications still reached unacceptably high rates even in highly experienced physicians.

## Discussion

In this in-depth contemporary analysis of the tfCAS learning curve, we demonstrated that this procedure continues to be subject to a lengthy learning curve with decreasing complication rates with increasing experience. Higher physician experience was associated with a decrease in rates of in-hospital stroke/death, in-hospital stroke/death/MI, 30-day mortality and in-hospital stroke/TIA in both symptomatic and asymptomatic patients. Additionally, the in-hospital stroke/death rate remained above 4% until 235 procedures in symptomatic patients and above 2% until 13 procedures in asymptomatic patients. However, these procedures may not represent physicians’ actual first procedures, but rather the first procedures recorded in the VQI registry.

Physician experience with tfCAS has been repeatedly shown to correlate with decreased post-operative stroke/death rate. In a study by Diethrich et al,^13^ there was a notable reduction in the rates of stroke and death, declining from 10.9% in the initial 110 cases to 6.2% in the subsequent 179 cases. Ahmadi et al found higher rates of neurologic complications post-tfCAS in the first 80 cases compared with the subsequent 240 cases.^14^ In an analysis of 200 tfCAS procedures, the initial 50 cases exhibited longer procedure durations and elevated stroke rates in comparison to the latter 150 cases.^15^ In a study based on CAPTURE2 that included 3388 tfCAS procedures by 459 physicians, a threshold of 72 procedures was needed to achieve 30-day stroke/death rates below 3% in asymptomatic non-octogenarian patients.^16^ Additionally, the Italian Consensus for Carotid Stenting/Stroke Prevention and Educational Awareness Diffusion ICCS/SPREAD guidelines suggest a cutoff of 75 procedures, of which 50 should be done as primary operator within a 2 year timespan to reach an acceptable risk of perioperative outcomes.^26^ In a separate study by Carrafiello et al that examined patients at high risk for carotid surgery, death/stroke rate was 8% in the first 50 patients vs 1.6% in the following 59 cases, although the results did not reach statistical significance.^27^ In a study analyzing the Pro-CAS German registry based on center experience, in-hospital stroke/death decreased from 5.9% in a center’s first 50 procedures to 4.5% in procedures 51-150, to 3.0% in procedures 151+ for both symptomatic and asymptomatic patients.^28^ These studies further reinforce the strong learning curve effect in the tfCAS procedure. Despite advancements in technology, the stroke/death rate remains unacceptably high with a persistent and lengthy learning curve.

Though tfCAS was first introduced as a potential alternative to CEA in high-risk patients, its higher perioperative stroke/death and other complication rates have hindered its acceptance as a substitute despite its minimally invasive nature.^1,3^ However, the introduction of TCAR offers a procedure that, although a little more invasive than tfCAS, demonstrates improved outcomes and a shorter learning curve.^5,6,7,29,30^ A previous study that analyzed the TCAR learning curve based on the VQI showed a short curve with in-hospital stroke/death rates of 1.5% and stroke/death/MI rates of 1.8% in physicians’ first 5 cases.^29^ In a separate study with combined symptomatic and asymptomatic patients, the 30-day stroke/death and stroke/death/MI rates were both 2.4% in physicians’ first 15 cases.^31^ In comparison, our current study showed a 5.2% in-hospital stroke/death rate and 5.8% in-hospital stroke/death/MI rate in symptomatic patients in the initial cases, and 2.1% and 2.6% respectively in asymptomatic early cases. This observation aligns with other studies^2,29–34^ that consistently reported lower peri-operative complications for TCAR compared to tfCAS.

According to several studies, 25-30% of perioperative stroke/death post-CAS occur after discharge.^18,19,20^ Similarly, two separate studies showed that 10-37% of strokes post-CEA occur after discharge; however, these were limited by small sample sizes or incomplete follow-up.^35,36^ Based on the National Surgical Quality Improvement Program (NSQIP), Fokkema et al showed that 40% of perioperative stroke/death post-CEA occur after discharge.^37^ As a result of the substantial post-discharge event rate, the European Stroke Organization suggested modified thresholds for in-hospital stroke/death rates to be 4% for symptomatic and 2% for asymptomatic patients, compared to the 6% for symptomatic and 3% for asymptomatic 30-day event rates.^21,22^ As the VQI has the most complete data for in-hospital stroke/death rates compared to 30-day events, we set thresholds of 4% for symptomatic and 2% for asymptomatic to define acceptable in-hospital stroke/death rates. In our study, stroke/death rate reached the cutoffs after 235 VQI procedures in symptomatic and 13 VQI procedures in asymptomatic patients. The greater proportion of low volume physicians after the recent CMS coverage expansion would lead to a great number of tfCAS being performed with unacceptable rates of perioperative stroke/death.

To gain further insight into our observed learning curve effect, we conducted a sensitivity analysis which revealed that physicians who performed a higher number of procedures had better outcomes from the start. One possible explanation is that doctors with superior initial outcomes were more motivated to keep performing tfCAS compared to those with worse initial outcomes. Moreover, mandatory reporting requirements for institutions and the mandate for institutional criteria and case review were put in place to ensure adequate quality in reporting complication rates, which may have influenced the results. With these mandates, physicians who have worse initial outcomes may be more likely to stop performing the procedure early on. Without these mandates and registry participation, physicians would not necessarily know their outcomes and would be less likely to stop performing tfCAS despite high complication rates. To assess the improvement per physician, we ran an additional analysis using the GEE model with clustering based on physician ID. That way, the GEE allowed us to isolate the effect of experience on each physician’s post-operative outcomes. The results remained statistically significant after controlling for that effect, validating the effect of experience on decreasing post-operative complications.

A fundamental limitation of our study is its retrospective nature. Initial cases used in our analysis likely do not represent physicians’ true initial procedures given that initial procedures may have been performed prior to participation in the VQI registry. The VQI was created from the Vascular Study Group of New England (VSGNE) which was expanded into a national database only in 2011. Since CAS was being performed widely within the New England region prior to 2005, the earliest recorded case in the VQI, our results do not represent the initial cases from surgeons in that region. Moreover, CAS was being performed widely throughout the country well before the expansion in 2011, so VQI results probably do not represent initial cases for many physicians. Thus, it is safe to conclude that the learning curve is underestimated.

Moreover, although VQI centers are routinely audited for completeness of patient entry, there is no routine auditing for source data accuracy. Nevertheless, the data on in-hospital mortality is validated.

## Conclusion

Post-operative complications after tfCAS decreased with increasing physician experience, showing a persistent lengthy learning curve consistent with previous reports. Given the fact that physicians’ early cases may not be included in the VQI, the learning curve was almost certainly underestimated. Nevertheless, tfCAS performed by physicians with low procedure counts showed significantly elevated complication rates. After the recent CMS coverage expansion for tfCAS, a significant number of physicians will enter the early stage of the learning curve, likely leading to increased post-operative complications.

## Data Availability

Data are available upon request for all VQI centers. R codes for data analysis are available upon request.

## Grant support

- This work in part was funded by NIH 5T35HL110843 fellowship award to “Strauss, S.”
- AS is supported by the Harvard-Longwood Research Training in Vascular Surgery NIH T32 Grant #5T32HL007734-29
- This work was conducted with support from Harvard Catalyst (NIH grant #UL1 TR002541)

## Conflicts of Interest

None

## Notes

### Competing Interest Statement

The authors have declared no competing interest.

### Author Declarations

This study was approved by the Beth Israel Deaconess Medical Center institutional IRB, informed consent for participation was waived due to the deidentified retrospective nature of the data.

